# Protocol for: Diagnostic trajectories of patients receiving unspecified psychosis-spectrum diagnoses – A retrospective, longitudinal, register-based study in a large UK secondary mental healthcare service from 2007–2024

**DOI:** 10.64898/2026.10.22.26366201

**Authors:** Eoin Gogarty, Cameron Watson, Maximin Lange, Ben Carter, Sameer Jauhar, Robin Murray, Robert Stewart, Matthew Broadbent, Samuel Swidzinski, Katherine Beck, Ana Rita Moura, Fiona Gaughran

## Abstract

**Background:** Unspecified psychosis-spectrum diagnoses (ICD code F29) are commonly used when the underlying disorder remains uncertain, particularly at first presentation. These diagnoses may persist or later be revised as the clinical picture evolves, yet little is known about their long-term trajectories or implications for treatment and service use in routine mental healthcare.

**Objectives:** To describe the usage and persistence of unspecified non-organic psychosis (F29) and other non-specific diagnoses, and the time to transition to a specific diagnosis. Secondary objectives are to describe temporal trends in their use, identify demographic and clinical characteristics associated with diagnostic persistence, and characterise treatment patterns, service utilisation, mortality, and discharge across diagnostic groups.

**Methods:** We will conduct a retrospective longitudinal cohort study using de-identified electronic health-record data from the South London and Maudsley NHS Foundation Trust (SLaM), accessed through the Clinical Record Interactive Search (CRIS) system, between 1 January 2007 and 31 December 2024. The study population will include patients with an ICD-10 psychosis-spectrum diagnosis (F2x.x), substance induced psychotic disorder (or F1x.5), bipolar disorder or affective disorder with documented psychotic symptoms(F32/3.3). Diagnoses will be classified as specific or non-specific according to their ICD-10 category. Primary analysis will examine time from first F29 diagnosis to first subsequent specific psychosis-spectrum or bipolar diagnosis using Cox proportional hazards regression, with discharge from SLaM, death and administrative end of follow-up treated as censoring events. Associations with diagnostic transition will be examined adjusting for age, sex, ethnicity, area-level deprivation, route of service entry, and calendar period. Secondary competing-risk analysis will estimate the cumulative incidence of diagnostic transition using the Aalen– Johansen estimator, treating discharge and death as competing events.

**Ethics and Dissemination:** CRIS operates under ethical approval from the South Central–Oxford C Research Ethics Committee (23/SC/0257). Project-specific approval was received from the CRIS Oversight Committee as Project 23-095. The study will be reported in accordance with the RECORD extension of the STROBE guidelines. Findings will be disseminated through peer-reviewed publication and will provide evidence on the longitudinal use and refinement of non-specific psychosis-spectrum diagnoses in routine secondary mental healthcare.

## Background

Psychotic disorders are clinically heterogeneous, and diagnosis assigned at first presentation may change as symptoms evolve and additional information becomes available. Diagnostic uncertainty is hence common in early psychosis, when clinicians may use non-specific diagnoses such as ICD-10 F29 (unspecified non-organic psychosis) before a more specific disorder can be established, with F29 intended principally as a provisional diagnosis [1–3]. Diagnosis informs treatment, prognosis and pathways through mental health services, while long-term outcomes may relate more closely to later diagnostic classification than to the diagnosis assigned at first presentation [4,5]. Crucially, there is no corresponding diagnosis-specific evidence-based treatment pathway for F29. By contrast, established diagnoses such as schizophrenia and bipolar disorder are linked to established clinical guidelines and treatment recommendations. Prolonged retention of an unspecified diagnosis may therefore have implications not only for diagnostic accuracy, but also for access to appropriately targeted treatment and longer-term care.

Diagnostic stability differs substantially across the psychosis spectrum. Schizophrenia and affective psychoses tend to show high longitudinal stability, whereas unspecified, brief, acute and other non-specific psychotic diagnoses are more likely to be revised [4,6–12]. Meta-analyses support this distinction, demonstrating lower stability for psychosis not otherwise specified than for schizophrenia and affective psychoses [6], and considerable diagnostic change following brief or acute and transient psychotic disorders [7,8]. Where diagnostic change occurs, transition towards schizophrenia-spectrum disorders is common, although transitions towards affective disorders also occur [6–8]. These patterns are further observed in prospective first-episode cohorts [4,9–11] and in patients with co-occurring substance-use disorders, where diagnostic stability has been reported to be highest for schizophrenia-spectrum diagnoses and lowest for unspecified diagnoses [12]. Longer-term studies further suggest that diagnostic revision can continue over many years and can cross psychotic and affective diagnostic boundaries [5,11].

Despite extensive evidence of diagnostic instability in psychosis, little is known about the longitudinal use of non-specific diagnoses within UK routine secondary mental healthcare. Existing evidence has predominantly come from first-episode cohorts, structured research assessments, specific short-duration psychotic disorders, or relatively small clinical samples [3,4,6–12]. Using longitudinal electronic health-record data from a large secondary mental healthcare provider, we therefore aim to describe the incidence and prevalence of non-specific psychosis-spectrum diagnoses, examine their persistence and transition to specific diagnoses, identify demographic and clinical characteristics associated with diagnostic trajectory, and describe treatment patterns, service utilisation, discharge and mortality according to diagnosis held during follow-up.

## Methods

### Aims and Objectives

This study investigates the incidence and prevalence of non-specific psychosis-spectrum diagnoses among patients presenting with psychotic symptoms in the context of a schizophrenia spectrum or affective disorder, and their demographic and clinical characteristics, diagnostic trajectory, and service use at a large secondary mental healthcare provider over an 18-year period.

#### Primary objective

To describe the persistence of non-specific psychosis-spectrum diagnoses and time to transition to a specific diagnosis.

#### Secondary objectives

1. To describe the distribution and temporal trends of first psychosis-spectrum diagnoses.
2. To describe patient characteristics associated with persistence of unspecified diagnoses.
3. To describe treatment patterns, service utilisation, mortality, and discharge from South London and Maudsley NHS Foundation Trust (SLaM) in relation to diagnosis.

### Study design

Retrospective longitudinal cohort study using routinely collected electronic health records.

### Database

The Clinical Record Interactive Search (CRIS) [13] contains de-identified electronic health record data from SLaM. CRIS operates under ethical approval from the South Central – Oxford C Research Ethics Committee (23/SC/0257), while this present study has project-specific approval from the CRIS Oversight Committee for Project 23-095. The observation period will be 1 January 2007 to 31 December 2024.

### Setting

SLaM is the largest specialist mental health NHS trust in the UK, providing secondary mental healthcare to over 1.3 million people across the South London boroughs of Lambeth, Southwark, Lewisham and Croydon. It delivers a broad range of inpatient, community and specialist mental health services.

### Inclusion and Exclusion Criteria

#### Inclusion criteria

Patients will be eligible if they have at least one qualifying ICD-10 psychosis-spectrum or bipolar/affective disorder diagnosis (F2x, F30, F31, F32.3, F33.3, F39, or F1x.5) during the observation period. Patients with bipolar disorder (F30/F31) and unspecified mood disorder (F39) will be included only where at least one distinct positive psychotic symptom is documented using the validated CRIS NLP applications (delusions, hallucinations (auditory, visual and other), paranoia, passivity, persecutory ideation, thought broadcast, thought insertion, and thought withdrawal). Behavioural and activation symptoms (e.g. aggression, agitation, arousal, hostility and irritability) will not contribute to this criterion.

For the primary analysis of diagnostic persistence, the index diagnosis will be F29 and must be the patient’s first qualifying diagnosis, with no earlier specific diagnosis.

#### Exclusion criteria

Patients who have opted out of their de-identified clinical records being included in in the deidentified database will not be included. People whose only recorded team in SLaM is the at-risk mental state (OASIS) service will be excluded from the analysis. In accordance with the CRIS security model, statistical cell sizes of fewer than 10 individuals will not be reported, to minimise the risk of inadvertent deanonymisation.

### Clinical Variables

#### Demographics

Age at first SLaM presentation (a proxy for age at illness onset; continuous, and categorised as <25, 25–34, 35–44, and ≥45 years), sex, self-reported ethnicity (ONS five-category), and Index of Multiple Deprivation (IMD) decile at first recorded address.

#### Diagnostic episodes

A diagnostic episode will be defined as a continuous period under a single primary ICD-10 psychosis-spectrum or affective disorder diagnosis; episodes will be carried forward across gaps in active SLaM contact for classification of the diagnosis held at landmarks, and a new episode will begin only on transition to a different psychosis-spectrum or bipolar code. An indicator of comorbid substance-use disorder (any non-psychotic F10– F19 code excluding F1x.5, recorded at or before the index date) will also be derived.

#### Medication Exposure

Medications will include antipsychotics (clozapine and non-clozapine, with long-acting injectables identified separately), mood stabilisers (lithium, valproate), antidepressants, and benzodiazepines. The date of first recorded mention with a documented dose will be taken as a proxy for treatment initiation.

#### Service entry

Service entry will be classified as inpatient or community according to the SLaM service to which the patient was first accepted at the index presentation.

#### Diagnostic Classification

##### Specific Diagnoses

Schizophrenia (F20), schizotypal disorder (F21), delusional disorder (F22), induced delusional disorder (F24), schizoaffective disorder (F25), bipolar disorder (F30/F31 with documented psychotic symptoms), and psychotic depression (F32.3/F33.3).

##### Non-specific Diagnoses

Unspecified non-organic psychosis (F29), other non-organic psychotic disorders (F28), unspecified mood (affective) disorder (with psychotic symptoms) (F39), acute and transient psychotic disorder (F23), and substance-induced psychotic disorder (F1x.5), which leave the underlying disorder undetermined.

The documented psychotic-symptom requirement will apply to cohort entry through bipolar disorder (F30/F31) or unspecified mood disorder (F39). For patients already included in the cohort, a subsequent F30/F31 diagnosis will count as a specific destination diagnosis irrespective of documented psychotic symptoms.

### Outcomes

#### Primary outcome

Time from first F29 diagnosis to first subsequent specific psychosis-spectrum or bipolar diagnosis. For bipolar disorder, documented psychotic symptoms will be required for cohort entry but not for classification as a subsequent destination diagnosis.

#### Secondary outcomes

- Primary ICD-10 code at first presentation and at each landmark.
- Proportion discharged from SLaM by each landmark, stratified by diagnosis at first presentation, together with diagnosis at discharge.
- Re-presentation to SLaM following discharge, including the timing and frequency of subsequent episodes of care.
- Service use between first presentation and each landmark (total bed days, number of distinct inpatient admissions, HTT days, PICU days, emergency department (ED) liaison contacts, Mental Health Act (MHA) detentions, and community mental health team (CMHT) contacts), per year active in SLaM.
- All-cause mortality, analysed as an exploratory comparison across diagnostic groups at first presentation and at each landmark.
- Treatment receipt (one antipsychotic; two or more distinct antipsychotics, including clozapine and long-acting injectable forms; clozapine; long-acting injectable (depot); lithium; valproate; each as ever prescribed with documented dose, and time to first recorded mention with documented dose).

Secondary outcomes will be assessed at Year 1 (12 months) and Year 4 (48 months) following first presentation. The Year 1 landmark will capture early diagnostic refinement and service use, while the Year 4 landmark will provide longer-term follow-up after most patients would be expected to have moved beyond time-limited Early Intervention services. In accordance with earlier work, the index date will be the date of the first qualifying diagnosis recorded in the structured CRIS/ePJS diagnosis field.

#### Descriptive Statistics

Descriptive statistics will be presented for all variables. Continuous variables will be reported as means with standard deviations or medians with interquartile ranges depending on their distribution. Categorical variables will be presented as frequencies and percentages. The study population will be described overall and by diagnostic category. Annual counts and proportions of first diagnoses in each diagnostic category will be reported across the observation period, and the proportion of patients active at each calendar year-end holding each diagnostic category will be described over time.

#### Primary Statistical Model

Time to diagnostic transition will be analysed using Cox proportional hazards regression. Follow-up will begin at the first F29 diagnosis and continue until diagnostic transition, discharge from SLaM, death, or 31 December 2024, whichever occurs first. Diagnostic transition will be treated as the event, while discharge, death and administrative end of follow-up will be treated as censoring events. Associations with diagnostic transition will be examined adjusting for age at first presentation, sex, ethnicity, Index of Multiple Deprivation decile, service entry, and calendar period of first diagnosis. Adjusted hazard ratios with 95% confidence intervals will be reported. The proportional hazards assumption will be assessed.

#### Secondary analyses

The cumulative incidence of diagnostic transition from first F29 diagnosis to a first subsequent specific psychosis-spectrum or bipolar diagnosis will be estimated using the Aalen–Johansen estimator, with discharge from SLaM and death treated as competing events. Re-presentation following discharge will be summarised descriptively.

At 1 and 4 years, patients in active SLaM contact, including those re-referred after an earlier discharge, will be classified according to the primary ICD-10 diagnosis held at the respective landmark. Discharge and mortality by each landmark will be assessed in the full cohort, while other outcomes will be assessed among patients active at the respective landmark.

Service utilisation outcomes, including numbers of inpatient admissions, bed days, Mental Health Act detentions and community contacts, will be analysed using negative binomial regression with an offset for duration of active SLaM contact. Binary outcomes, including treatment receipt, will be analysed using logistic regression. Time to first recorded prescriptions will be examined using competing-risks methods. Adjusted analyses will use the same covariates as the primary analysis.

For each specific destination diagnosis, including schizophrenia, schizoaffective disorder and bipolar disorder, the number and proportion of patients transitioning will be reported, together with the median (IQR) observed time from first F29 diagnosis to transition among those who transition. Median time to first discharge will also be reported, with results additionally summarised by sex and ethnicity.

#### Subgroup analyses

Subgroup analyses of the primary outcome will be conducted by sex, ethnicity, age group, and comorbid substance-use disorder. The association with inpatient admission within six months of first presentation will be examined in a six-month landmark analysis restricted to patients who remain under follow-up and have not experienced diagnostic transition by six months.

#### Sensitivity analyses

We will assess the robustness of the primary and secondary findings to differences in follow-up, service context, symptom ascertainment, and service-use intensity and continuity as indirect proxies for illness severity. Analyses will be repeated: (1) restricting the cohort to patients with at least 12 months of active SLaM contact after the index diagnosis; (2) restricting the primary analysis to patients with ongoing SLaM contact at Year 4; (3) restricting the cohort to patients with at least 12 clinical contacts in each year of active follow-up; (4) restricting the cohort to patients with at least two inpatient admissions; (5) excluding patients whose first diagnosis was recorded within an OASIS at-risk mental state service at the Year 1 landmark, while retaining those still receiving psychosis-spectrum care at Year 4, reflecting the time-limited nature of OASIS care; (6) restricting the cohort to patients with at least one documented positive psychotic symptom at or before the index diagnosis, thereby requiring symptom corroboration across all diagnostic groups; and (7) restricting analyses to patients who were never discharged from SLaM during follow-up, to reduce the influence of unobserved diagnostic changes outside SLaM. Analyses conditioning on post-index service use, continued service contact or later retention will be interpreted cautiously because they describe selected patient subgroups rather than the full cohort.

#### Handling of missing data

Multiple imputation by chained equations will be conducted as a sensitivity analysis for covariates with more than 10% missingness; the imputation model will include the event indicator, follow-up time, and all analysis covariates. Missing ethnicity will additionally be reported as an explicit category in descriptive tables.

#### Software and Reporting

All analyses will be conducted in Python. We will report in accordance with the RECORD extension of the STROBE guidelines [15] and all analysis code will be made publicly available.

## Limitations

We note that differences in outcomes between diagnostic groups may reflect both diagnostic practice and underlying clinical heterogeneity and should therefore not be interpreted as causal effects of the diagnostic label itself. In particular, access to clozapine and mood stabilisers is partly determined by diagnosis, such that lower treatment receipt among patients with F29 may reflect prescribing pathways. Some misclassification of OASIS involvement is also possible, as service location might be missing from records. Diagnostic classification will be based on the primary ICD-10 code recorded on the ePJS summary page, which although it may not always capture the clinician’s current working formulation, represents the diagnosis available operationally to receiving clinicians. Treatment ascertainment will be limited to pharmacological treatment, and only lithium and valproate will be included among mood stabilisers; other agents such as lamotrigine and carbamazepine, as well as psychological therapies and ECT, will not be captured. Diagnostic trajectories are restricted to the specified psychosis-spectrum and bipolar codes, meaning that non-psychotic diagnoses occurring before, between or after psychotic episodes will not be observed. Our findings will reflect the diagnostic practices, service configuration and catchment of a single south London NHS Trust and may not generalise to other settings; practice may additionally have changed over the 18-year observation period, including through changes in early intervention provision. Finally, the first F29 diagnosis observed within the Trust may not represent the patient’s first-ever F29 diagnosis because earlier care or treatment elsewhere is unobserved, resulting in potential left truncation. Service-use intensity and continuity will be examined as indirect proxies for severity.

## Data Availability

All analysis code will be made publicly available after study conclusion

